# Cost analysis of a nationwide typhoid conjugate vaccine campaign in Burkina Faso

**DOI:** 10.64898/2026.04.17.26350856

**Authors:** Jean-Louis Koulidiati, L Robert Zoma, I Eric Nebié, Yameogo Soumaila, Ouedraogo Christelle Neya, Joël Arthur Kiendrébéogo, Fréderic Debellut

## Abstract

**Background:** In Burkina Faso, typhoid fever remains a major public health concern, with a high incidence among children younger than 15 years of age. To address this burden, the country introduced typhoid conjugate vaccine in January 2025 through a national vaccination campaign reaching children aged 9 months to 14 years.

This study aimed to estimate the cost of typhoid conjugate vaccine delivery during the national campaign and to identify the main cost drivers across different administrative levels.

**Methods:** We conducted a cross-sectional, retrospective costing study using a microcosting approach from the government perspective. We collected data from fifty health facilities, eight health districts, five health regions, and the national level. Financial and economic costs were estimated for each level, excluding vaccine and syringe costs. All costs were converted to 2024 USD using the official exchange rate.

**Findings:** Vaccinators administered a total of 10.5 million typhoid conjugate vaccine doses. The average financial cost per dose was $0.47 (95% CI: $0.39–$0.51), and the economic cost was $2.16 (95% CI: $1.71–$2.56). Human resources and per diem payments were the main contributors to costs. Costs varied by geography, delivery strategy, and security context, with higher costs observed in rural and conflict-affected areas. The mobile–temporary posts strategy had the highest economic cost per dose ($2.02; 95% CI: $1.64–$2.40), while the fixed strategy had the highest financial cost per dose ($0.41; 95% CI: ($0.32–$0.49).

**Conclusion:** The financial cost per dose remained within Gavi, the Vaccine Alliance’s operational support range. The observed cost variations highlight the need for targeted funding and enhanced logistical support to ensure equitable access, particularly in rural and insecure areas. This study provides evidence to inform future vaccination campaigns and supports decision-making for typhoid conjugate vaccine introduction in other countries in the region.

## Introduction

Typhoid fever remains a significant public health challenge globally, with more than 7 million cases and 93,000 deaths estimated in 2021 [1]. The burden of disease is disproportionately high in South Asia and sub-Saharan Africa, where poor sanitation and lack of access to clean water—main transmission routes of the disease—are common [2]. In sub-Saharan Africa, more than three million infections and 33,000 deaths occur annually, with incidence rates reaching up to 383 cases per 100,000 person-years in some countries [3,4]. The region has also seen an increase in typhoid outbreaks, such as the 2023 outbreak in the Democratic Republic of Congo that recorded nearly 1,200 suspected cases, with typhoid intestinal perforations occurring in 3.7% of cases [5].

In response to this persistent threat, the World Health Organization (WHO) recommended the introduction of the typhoid conjugate vaccine (TCV) in endemic countries in 2018. Two vaccines—Typbar TCV® and TYPHIBEV®—are currently available for Gavi-eligible countries in liquid form in five-dose vials. Despite the high disease burden, only five African countries have introduced TCV to date [7].

Burkina Faso faces a substantial typhoid fever burden. Surveillance conducted in 2024 revealed an incidence rate of 133 cases per 100,000 person-years, with rates reaching up to 234 per 100,000 person-years among children aged 2 to 4 years. Children younger than 15 years, who constitute more than half of the population, accounted for 88% of reported cases [8]. In 2023, the country recorded over 120,000 suspected cases [9]. Progress has been made in disease surveillance, including the establishment of two sentinel sites under the Typhoid Surveillance in Africa Program (TSAP) [10,11], which enabled hospital-based monitoring and blood-culture confirmation. Also, in January 2025, Burkina Faso launched a national TCV campaign targeting children aged 9 months to 14 years old, reaching more than ten million recipients [12]. Following this campaign, the Ministry of Health integrated TCV into its routine immunization program.

Understanding the costs associated with vaccine delivery is essential for informed decision-making. However, cost data on TCV introduction, particularly in Francophone Africa, remain limited. This study addresses this gap by estimating the costs of TCV delivery during the campaign in Burkina Faso, providing evidence to support future planning and regional vaccine introduction efforts.

## 2. Methods

### Study design

We conducted a retrospective cross-sectional costing study using a micro-costing approach to estimate both financial and economic costs associated with the national TCV campaign in Burkina Faso. Financial costs included direct expenditures incurred during the campaign, while economic costs encompassed the value of all resources utilized, including volunteer time and donated materials, regardless of financial transactions [13]. The analysis adopted the perspective of the Ministry of Health, covering the entire campaign period—from preparatory activities to post-campaign review.

### Study sites and sampling

Burkina Faso’s health system is a hierarchical pyramid structure, comprised of 13 health regions, 70 health districts, and approximately 2,670 Health and Social Promotion Centers (CSPS), which serve as the primary level of care and the first point of contact for communities. All CSPS involved in the national vaccination campaign formed the sampling frame. We used a stratified random sampling approach and the EPIC Sample Design Optimizer (SDO) tool [14] to randomly select 5 regions, 8 districts, and 50 CSPS for inclusion. The 50 CSPS from 5 regions were chosen based on budgetary constraints. We stratified the sample based on urban versus rural location.

### Data collection

Data collection took place in March 2025, two months after the campaign ended, to reduce recall bias. Prior to fieldwork, data collectors received training on standardized questionnaires. They conducted visits at selected CSPS, district health offices, regional directorates, and the central level. Due to security concerns, we replaced five inaccessible CSPS with alternative facilities located in the same area and of similar size.

Enumerators collected primary data through interviews with key informants, including head nurses, Expanded Programme on Immunization (EPI) focal points, district and regional health managers, and staff from the Directorate of Vaccination Prevention (at the central level). Tailored questionnaires for each level captured information on campaign activities and cost categories as described in Table 1.

**Table 1.**
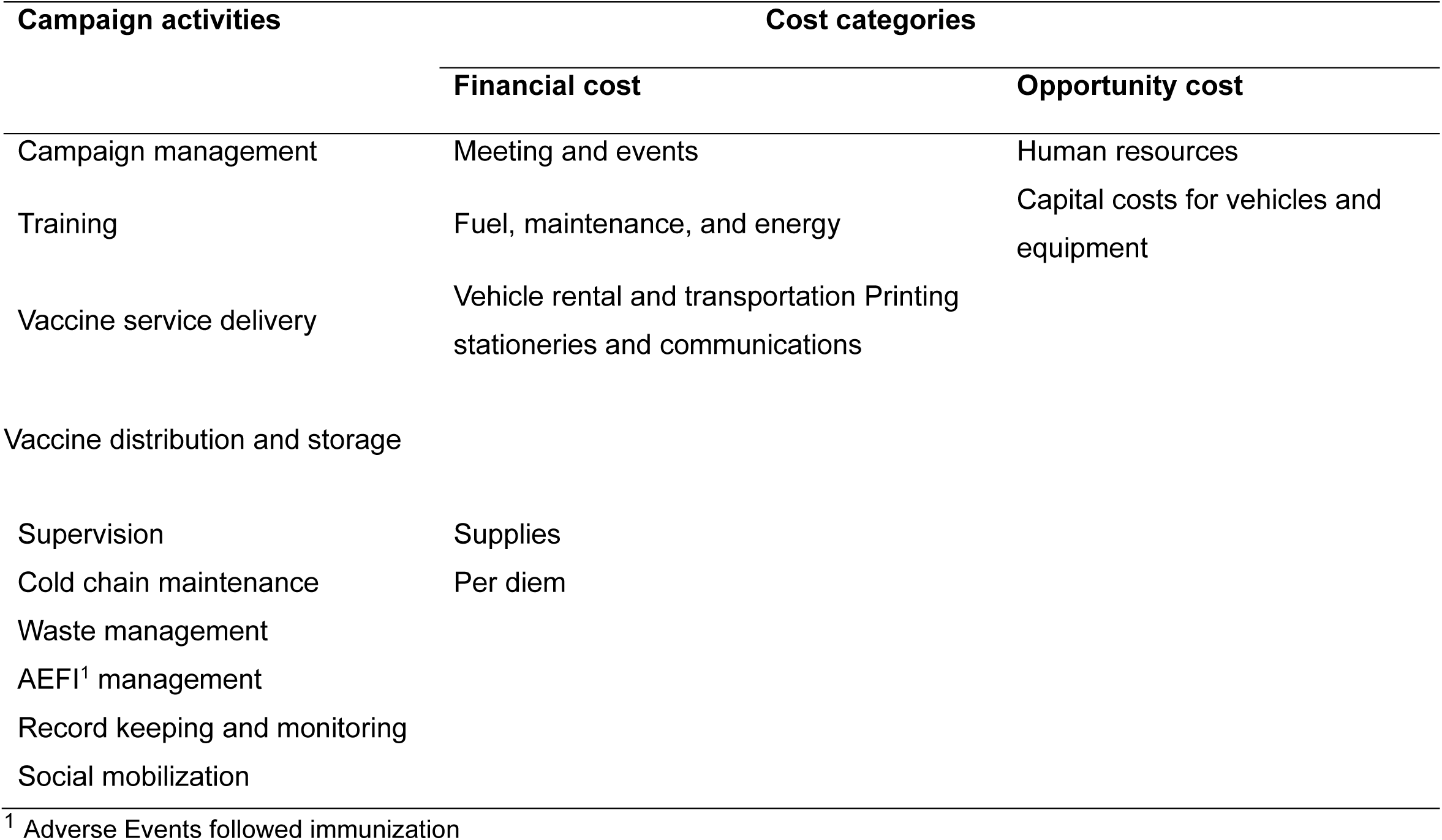
Campaign activities and cost categories.

Secondary data focused on unit prices, such as salary scales for campaign personnel and replacement costs for equipment and vehicles, sourced from government and international organizations.

### Data analysis

At each administrative level, we estimated the financial and economic costs of vaccine delivery for the campaign. We multiplied resource quantities by unit prices or opportunity costs to estimate activity-specific costs. Additionally, we calculated delivery costs by campaign activity and cost category excluding vaccine procurement and certain consumables (e.g., auto-disable syringes, safety boxes). Then, we aggregated costs per dose for each administrative level [15,16,17].

At the CSPS level, we calculated weighted average costs to obtain overall cost estimates. Furthermore, we calculated costs per dose by administrative level, delivery strategy, and geographic setting (urban/rural; conflict-affected/non-affected).

We collected all costs in West African CFA francs and converted them to 2024 USD using the World Bank exchange rate (1 USD = 606.35 F CFA) [18]. We annualized capital costs using replacement values, expected useful life, and a 3% discount rate [17]. We performed data analysis using Stata version 14 (StataCorp, TX, USA).

### Ethical considerations

Burkina Faso’s Health Research Ethics Committee granted ethical approval (Deliberation No. 2025-02-035) and the Ministry of Health approved the study. We secured oral informed consent from all participants.

## 3. Results

### Description of the study sample characteristics

The study sample comprised 50 CSPS, 8 health districts, 5 regions, and the central level. Of the fifty selected CSPS, 74% were in rural areas and 28% were in conflict-affected zones. Three CSPS (6%) also administered 212 doses of HPV and fourteen doses of MR vaccines in addition to TCV, representing less than 1% of total doses. We excluded these doses from the analysis.

Most CSPS (98%) implemented all three delivery strategies during the campaign. The mobile and temporary posts strategy—conducted in schools, markets, churches/mosques —was the most utilized, accounting for 50% of doses administered. Outreach at regular posts contributed 35%, while the fixed strategy represented 15%.

### Campaign costs

Table 2 presents the financial and economic costs incurred across administrative levels. At the CSPS level, the weighted average financial cost was $2,164, and the economic cost was $12,054. Service delivery accounted for the largest share of economic costs (41%), followed by social mobilization (24%). Per diems represented 89% of financial costs, while human resources contributed 81% of economic costs. All the study CSPS administered a total of 439,601 TCV doses, with an average of 8,792 doses per facility. The financial cost per dose was $0.39 (95% CI: $0.31–$0.40), and the economic cost per dose was $1.90 (95% CI: $1.62–$2.18).

**Table 2.**
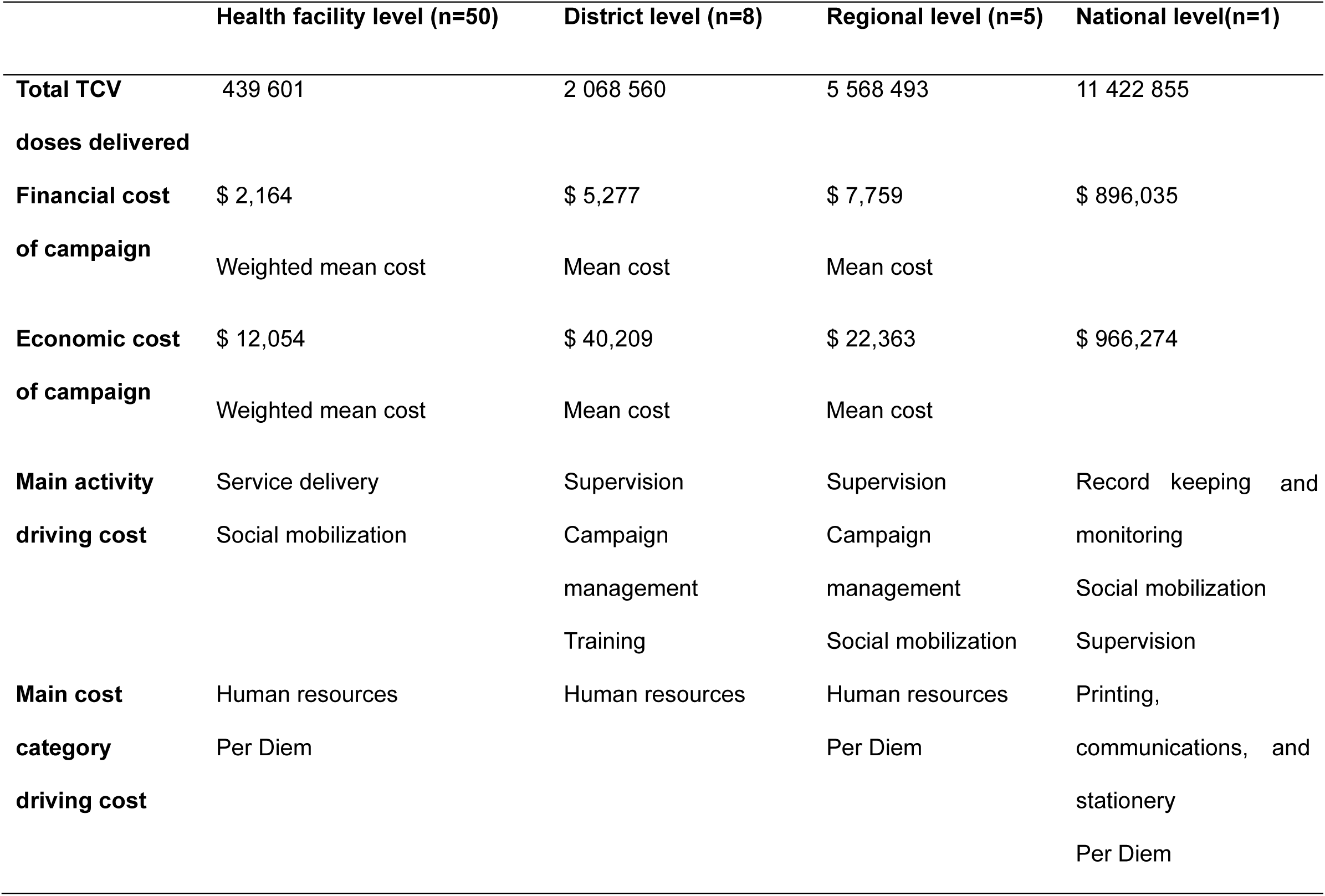
Number of doses delivered and campaign costs.

**Table 3.** Cost per TCV dose delivered.

|  | Health facility<br>level | District level | Regional level | National<br>level | All levels |
| --- | --- | --- | --- | --- | --- |
| Mean financial | \$0.35 | \$0.02 | \$0.01 | \$0.09 | **\$0.47** |
| cost (95% CI) | (\$0.31; \$0.40) | (\$0.01; \$0.03) | (\$0.00; \$0.01) | | **(\$0.39; \$0.51)** |
| Mean | \$1.90 | \$0.15 | \$0.02 | \$0.09 | **\$2.16** |
| economic cost<br>(95% CI) | (\$1.62; \$2.18) | (\$0.01; \$0.28) | (\$0.01; \$0.03) | | **(\$1.71; \$2.56)** |

At the district level, the average financial cost was $5,277, and the average economic cost was $40,209. Supervision and campaign management were the activities with the highest shares of both financial and economic costs. Human resources accounted for 85% of economic costs. Vaccinators administered 2,068,560 doses across the eight districts, resulting in a financial cost per dose of $0.02 (95% CI: $0.01–$0.03) and an economic cost per dose of $0.15 (95% CI: $0.01–$0.28).

At the regional level, the average financial cost was $7,759, and the average economic cost was $22,363. Supervision and campaign management were again the main activities driving costs, contributing 48% and 22% of economic costs, respectively. Human resources (61%) and per diems (19%) were the main cost categories. A total of 5,568,493 doses were administered in the five regions, with a financial cost per dose of $0.01 (95% CI: $0.00–$0.01) and an economic cost per dose of $0.02 (95% CI: $0.01–$0.03).

At the central level, the financial cost was $893,860 and the economic cost was $964,099. Data management and social mobilization were the main activities driving costs. Printing, communications, and stationery accounted for 65% of financial costs and 61% of economic costs, followed by per diems (15% and 14%, respectively). Nationally, 10,552,806 doses were administered, with both financial and economic costs per dose estimated at $0.09.

Across all levels, the average financial cost per TCV dose was $0.47 (95% CI: $0.39–$0.51), and the economic cost was $2.16 (95% CI: $1.71–$2.56).

### Cost distribution by activity and cost category

Figure 1 illustrates the breakdown of economic cost per dose by activity and cost category. Vaccine administration and social mobilization contributed $0.79 (37%) and $0.47 (22%) of the $2.16 economic cost per dose, respectively. Human resources accounted for $1.69 (78%) and per diems for $0.32 (15%).

**Figure 1.**
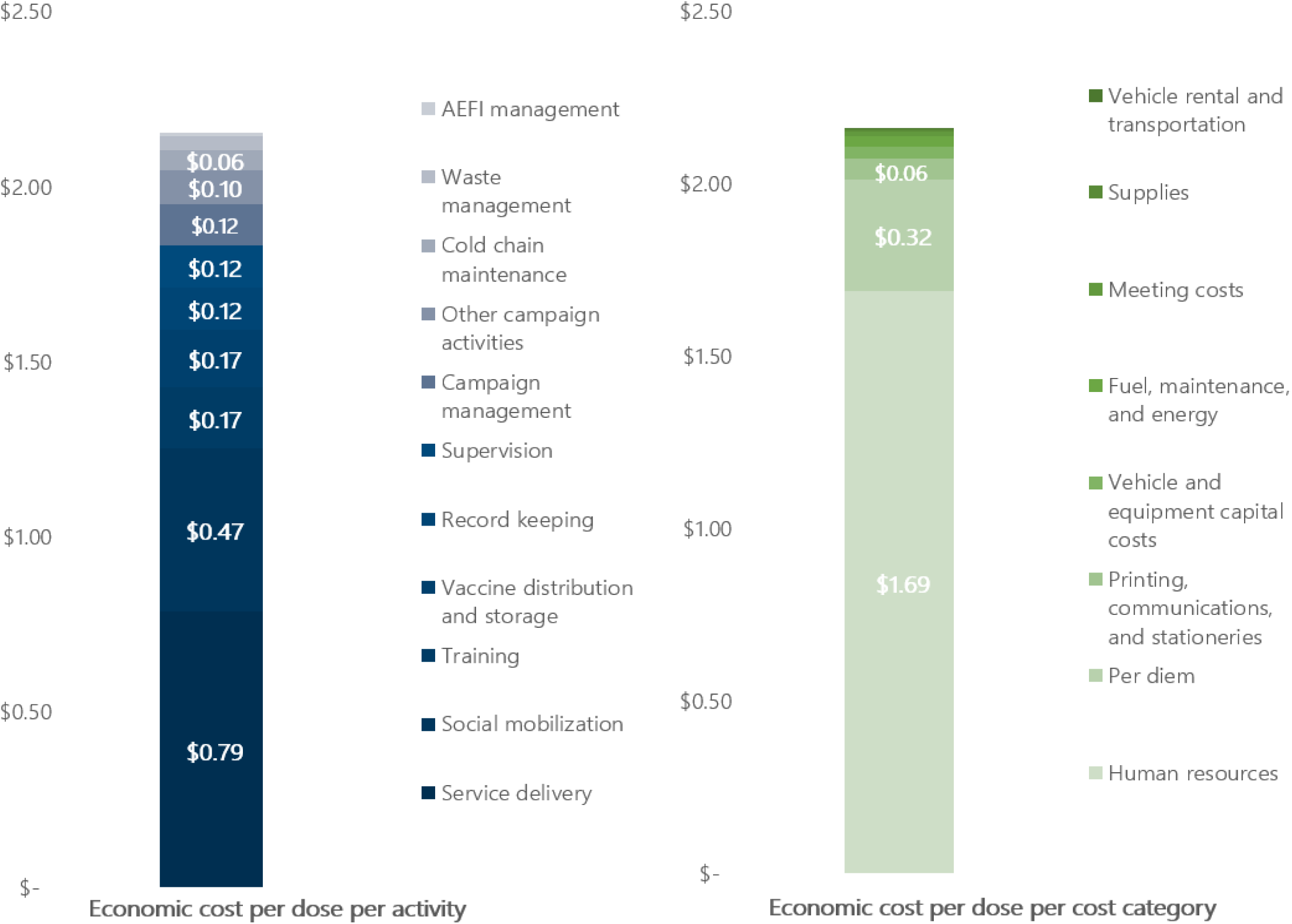
Economic cost distribution by activity and cost category.

### Cost variation by geography, delivery strategy, and security context

Table 4 shows cost variations by CSPS location, security status, and delivery strategy. Financial cost per dose was higher in rural areas ($0.40; 95% CI: $0.34–$0.45) than urban areas. The fixed strategy had the highest financial cost per dose ($0.41; 95% CI: $0.32–$0.49), while the mobile strategy had the highest economic cost ($2.02; 95% CI: $1.64–$2.40). CSPS in conflict-affected zones had a higher financial cost per dose ($0.39; 95% CI: $0.34–$0.44) than those in non-conflict-affected areas.

**Table 4.**
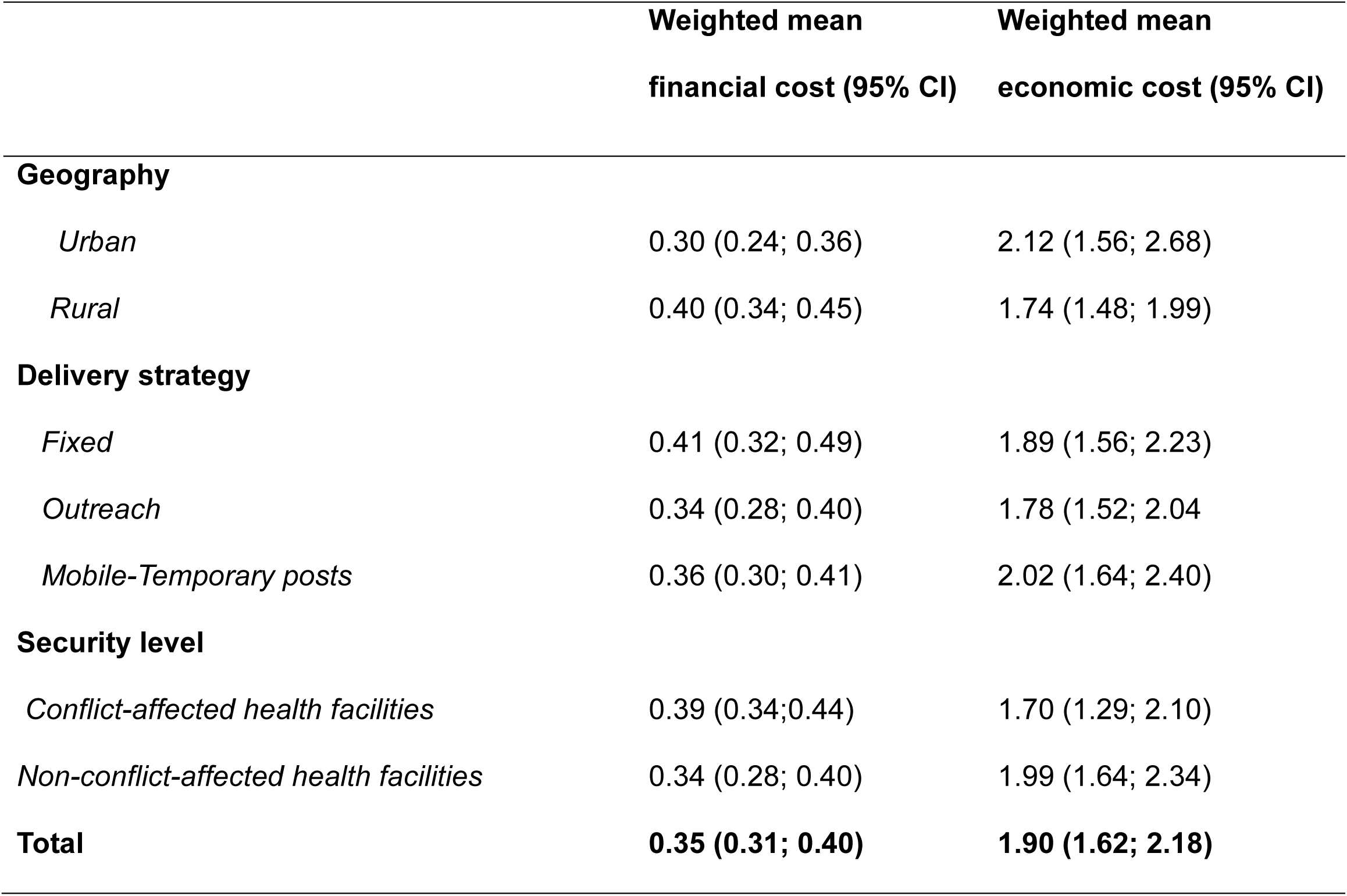
Cost per TCV dose by geography, delivery strategy, and security level (2024 USD)

|  | **Weighted mean**<br>**financial cost (95% CI)** | **Weighted mean**<br>**economic cost (95% CI)** |
| --- | --- | --- |
| **Geography** |  |  |
| *Urban* | 0.30 (0.24; 0.36) | 2.12 (1.56; 2.68) |
| *Rural* | 0.40 (0.34; 0.45) | 1.74 (1.48; 1.99) |
| **Delivery strategy** |  |  |
| *Fixed* | 0.41 (0.32; 0.49) | 1.89 (1.56; 2.23) |
| *Outreach* | 0.34 (0.28; 0.40) | 1.78 (1.52; 2.04) |
| *Mobile-Temporary posts* | 0.36 (0.30; 0.41) | 2.02 (1.64; 2.40) |
| **Security level** |  |  |
| *Conflict-affected health facilities* | 0.39 (0.34;0.44) | 1.70 (1.29; 2.10) |
| *Non-conflict-affected health facilities* | 0.34 (0.28; 0.40) | 1.99 (1.64; 2.34) |
| **Total** | **0.35 (0.31; 0.40)** | **1.90 (1.62; 2.18)** |

### Regional disparities and dose-cost relationship

Among the five regions, the East region had the highest financial cost per dose ($0.63), while the North region had the lowest ($0.39). The Hauts-Bassins region recorded the lowest economic cost per dose ($1.94), whereas the East had the highest ($2.53) (Figure 2).

**Figure 2.**
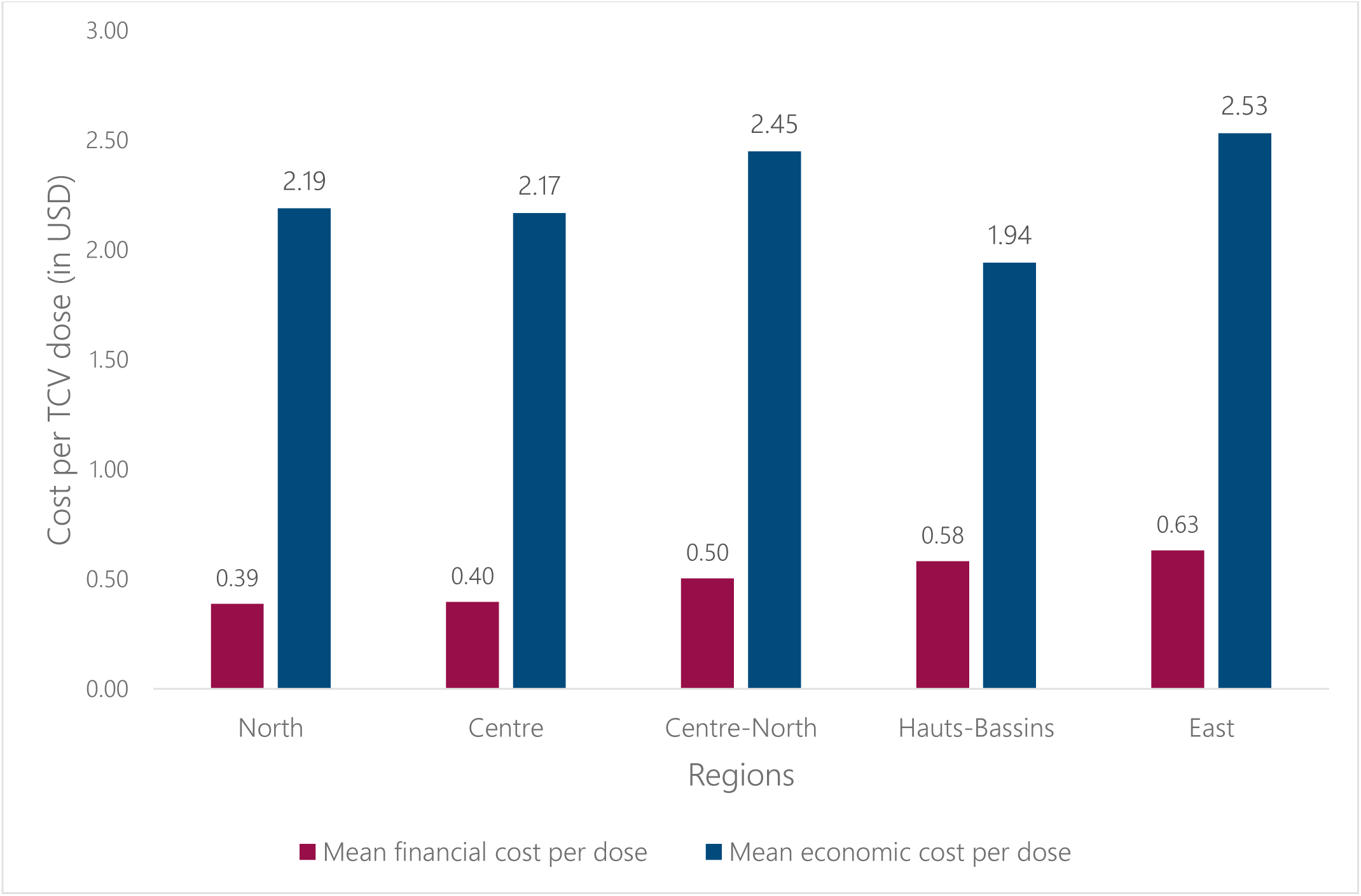
Financial and economic cost per dose by region.

Figure 3 shows the relationship between the number of doses administered and the associated costs at the CSPS level. The financial cost per dose decreased as the number of doses administered increased.

**Figure 3.**
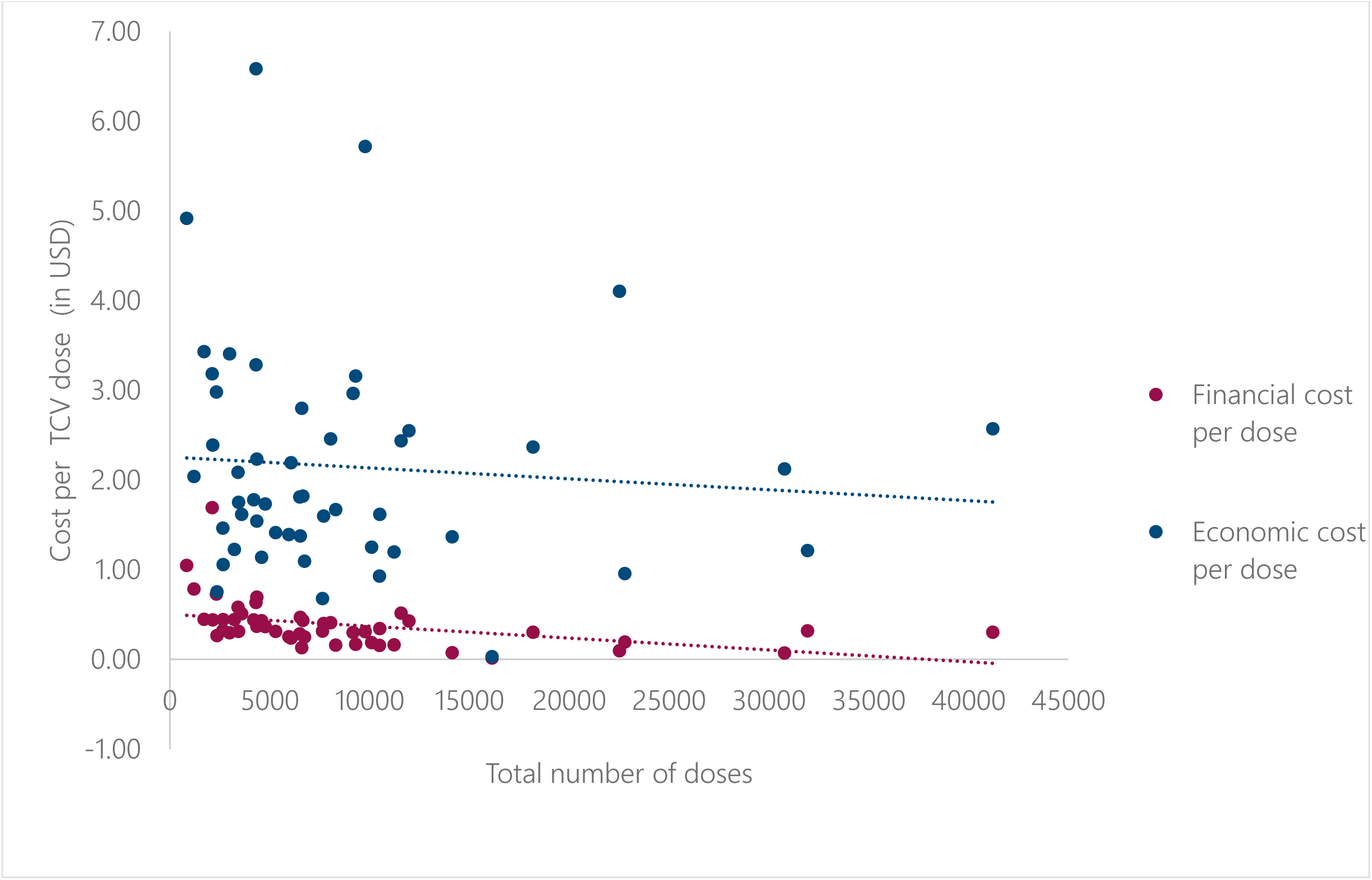
Relationship between costs and volume delivered at health facility level.

## 4. Discussion

This study provides a comprehensive assessment of costs associated with the national TCV campaign in Burkina Faso, encompassing all levels of the health system. The findings reveal substantial variability in cost per dose across administrative levels, geographic settings, and delivery strategies, offering critical insights for future immunization planning.

The average financial cost per dose of $0.47 observed in this study is lower than estimates from similar campaigns in other low- and middle-income countries. For instance, a typhoid outbreak response (TCV campaign) in Zimbabwe in 2019 reported a financial cost of $0.79 per dose [19], while Malawi’s 2023 integrated campaign yielded a financial cost per dose of $0.49 per dose [20]. The lower cost in Burkina Faso may be attributed to the strategic use of existing infrastructure and the implementation of multiple delivery approaches, which enhanced operational efficiency. Importantly, the financial cost per dose remains within the operational support range provided by Gavi ($0.45–$0.65) [21], suggesting that the campaign was financially viable under current donor frameworks. Nevertheless, the observed cost variability underscores the importance of context-specific budgeting and resource allocation.

A key finding is the predominance of human resources and per diems as cost drivers. Human resources accounted for the majority of economic costs, while per diems dominated financial costs. This pattern aligns with previous studies conducted in similar settings [21,22,23], where labor-intensive activities significantly influence immunization costs. Vaccine administration emerged as the main activity contributing to economic costs, followed by social mobilization. The latter’s prominence is unique compared to other campaign cost studies [19,20,22]. This reflects the substantial investment made during the campaign to address vaccine hesitancy and engage communities, political leaders, and education stakeholders. This emphasis on communication and outreach is particularly relevant given the increasing skepticism toward vaccines in various contexts.

At the central level, data management was the most resource-intensive activity, driven largely by the printing of over eleven million vaccination cards. This finding highlights the need to explore cost-saving alternatives, such as digital data collection and reporting systems, which could streamline operations and reduce expenditures.

Furthermore, the analysis also revealed that the fixed delivery strategy incurred the highest financial cost per dose, likely due to the lower volume of doses administered through this approach. This suggests the presence of economies of scale [22], where higher throughput leads to reduced unit costs. Conversely, the economic cost per dose for the fixed strategy was lower than for mobile and outreach strategies, possibly due to reduced reliance on additional human resources.

Regional disparities in cost were notable, with the East, Centre-North, and North regions—areas affected by insecurity—recording the highest economic costs. These elevated costs are probably due to increased logistical challenges and the need for additional personnel to ensure safe and effective vaccine delivery [22]. The higher opportunity costs in these regions further contribute to the overall economic burden. These findings emphasize the importance of tailoring campaign strategies and resource allocation to the specific needs of vulnerable and hard-to-reach populations.

The inverse relationship between the number of doses administered and financial cost per dose at the CSPS level further supports the existence of economies of scale [22]. Facilities administering larger volumes of vaccines achieved lower per-dose costs, reinforcing the value of optimizing delivery efficiency in future campaigns.

### Policy implications

The findings of this study have important implications for immunization policy and strategic planning. First, the high costs observed in rural and conflict-affected areas underscore the need for targeted financial and logistical support to ensure equitable vaccine access during campaigns. Tailored resource allocation strategies are essential to address geographic and security-related disparities.

Second, the substantial contribution of human resources to overall economic costs highlight the importance of investing in workforce development. Policymakers should prioritize training, retention, and efficient deployment of health personnel. Strengthening Burkina Faso’s community health strategy [24] could be instrumental in building a cadre of trained, community-based health workers capable of delivering essential services, including vaccination.

Third, the study identified data management—particularly at the central level—as a major cost driver. Transitioning to digital health information systems for real-time monitoring and evaluation could improve operational efficiency and reduce expenditures associated with printing and manual data handling [25].

### Study limitations

This study has several limitations. First, the retrospective nature of data collection may introduce recall bias or incomplete reporting. However, conducting fieldwork within two months of the campaign likely minimized this risk. Second, the cost analysis did not include depreciation of long-term infrastructure, which may lead to underestimation of total economic costs. Third, economic costs included estimates for volunteer time, but these may have been underreported due to reliance on self-reported data and lack of standardized valuation methods for non-financial contributions. Finally, due to insecurity, five CSPS were replaced with alternatives. This substitution may have introduced selection bias, but we selected facilities likely close in cost structure or campaign implementation to the replacing ones to reduce this bias.

## 5. Conclusion

This study provides a detailed estimation of the financial and economic costs of Burkina Faso’s national TCV campaign. The average financial cost per dose remained within the operational support range provided by Gavi, indicating financial feasibility. Human resources and per diems were the major cost drivers. Geographic and contextual variations in cost highlight the need for adaptive planning and targeted support. These findings offer valuable evidence to inform future vaccination campaigns and support TCV introduction in other countries facing similar public health challenges.

## Data Availability

All data produced in the present study are available upon reasonable request to the authors

